# Association Between Residence in Flood-Prone Areas and Incomplete Childhood Vaccination Coverage in Bangladesh

**DOI:** 10.64898/2026.02.11.26346067

**Authors:** Emilie Schwarz, Anna Dimitrova, Francois Rerolle, Tarik Benmarhnia, Kévin Jean

## Abstract

**Introduction:** Flooding events, which are strongly linked to climate change and variability, have the potential to disrupt communities and health systems. Vaccination, a highly effective public health intervention, plays a pivotal role in preventing numerous deaths annually, particularly among children. However, the relationship between exposure to flood events and early childhood vaccination remains unexplored.

**Methods:** This study utilizes validated flood exposure data from the Global Flood Database (GFD) and five waves of nationally representative survey data regarding the vaccination history of children under 3 years from the Bangladesh Demographic and Health Surveys (DHS) collected between 2004 and 2018. Using the geographical coordinates of each surveyed household grouping and matching it with the spatially resolved GFD data, we determined whether children reside in a flood-prone area. We then used Generalized Estimating Equations, accounting for geographical clustering and including an inverse probability of treatment weighting (IPTW) for covariate balance, to assess the relationship between living in a flood-prone area and incomplete vaccination. Incomplete vaccination was defined as having missed at least one does of the four World Health Organization-recommended childhood vaccines: Tuberculosis (BCG), Diphtheria-Tetanus-Pertussis (DTP), Polio (OPV), and Measles (MCV). The sensitivity of the association measure to various definitions of exposure was explored.

**Results:** Our sample included 13,649 children, of which 16% resided in flood-prone areas and 84% were fully vaccinated. Our findings indicate that children living in a flood-prone area have a higher risk of not receiving the DTP vaccine (PR = 1.24, CI: 1.05-1.46), the BCG vaccine (PR = 1.27, 1.08-1.50), and the OPV vaccine (PR = 1.24, CI: 1.06-1.46). The direction of the relationship remained but the effect became non-significant when looking at the MCV vaccine alone (PR = 1.06, CI: 0.90-1.25), as well as the four vaccines together (PR = 1.05, CI: 0.90, 1.22).

**Interpretation:** These findings hold critical implications for disaster management protocols, emphasizing the need to ensure uninterrupted access to routine healthcare services during floods, which are becoming increasingly more common due to climate change. Considering that populations in low- and middle-income countries are disproportionately impacted by extreme climate events such as floods and that preventable infectious diseases remain among the leading causes of child death in these regions, ensuring access to essential vaccines is critical for safeguarding public health.

## Introduction

According to the World Health Organization (WHO), climate change is the greatest threat to human health and directly contributes to a growing number of humanitarian emergencies.^1^ Extreme flood events, defined by the WHO as instances where excess water submerges normally dry areas, are the most common type of natural disaster worldwide.^2^ An estimated 255–290 million people were directly affected by floods between 2000 and 2018, and this number is projected to increase in the coming years as flood events become more frequent and severe.^3^

Extensive research has examined the direct and indirect impacts of floods on public health.^4–10^ There is evidence that flooding is associated with an increased risk of mortality, worsened mental health outcomes, and elevated prevalence of asthma and infectious diseases.^6,9,10^ A study published in 2023 also highlighted the long-term impacts on child mortality associated with living in flood-prone areas^10^. Besides these direct health impacts, extreme precipitation events may also indirectly impact population health by damaging infrastructure, preventing access to healthcare facilities, and limiting the capacity of healthcare systems to provide comprehensive care.

Vaccination is among the most impactful and cost-effective public health interventions globally, specifically in early childhood. Indeed, most vaccine-prevented deaths are among children under five.^11^ Yearly, an estimated 4 million deaths are prevented worldwide due to early childhood immunization. By 2030, the measles vaccine alone is estimated to save 19 million lives.^12^ Even though early childhood immunization is known to be remarkably effective, gaps still exist in vaccination coverage globally, especially in the post-COVID-19 context.^13–15^

Natural disasters, including extreme weather events, may hinder access to vaccination. Previous studies have identified the following pathways through which extreme weather events could theoretically influence vaccination: by displacing populations, damaging critical infrastructure, preventing access to routine services, disrupting supply chains, and diverting resources to other sectors.^16–19^ However, to date, there have been no empirical studies linking incomplete childhood immunization to flood exposure.

Bangladesh is consistently ranked among the countries most impacted by climate change, partly due to its network of 230 rivers and low-lying topography which pose a considerable risk of flooding.^20,21^ In recent years, an increase in the frequency of floods has been observed in the region, resulting in the loss of hundreds of lives and the displacement of millions.^21^ In 1979, the Expanded Program on Immunization (EPI) was launched in Bangladesh with the aim of providing access to essential vaccines as recommended by the WHO. Despite the notable improvements in the immunization rates since then, inequities in coverage remain. The high risk of flooding in Bangladesh and the presence of a government-operated vaccination system, provide a context in which the association between floods and early childhood immunization can be studied. We therefore assessed whether living in a flood-prone area would lead to a decrease in the probability of vaccination among children in Bangladesh.

## Methods

### Data

Vaccine information was retrieved from the Bangladesh Demographic and Health Surveys (DHS). DHS surveys have been routinely administered in over 90 low- and middle-income countries (LMICs) and are a source of information on various aspects of population health. The surveys are designed using a two-stage sampling procedure, which ensures their representativeness both at the national and subnational levels and for urban and rural areas^22,23^. Recent DHS waves include global positioning system (GPS) information (latitude and longitude) for each primary sampling unit (PSU). A PSU is defined as a city block in an urban area and a village in a rural area. We extracted information from the five most recent survey waves for Bangladesh which contain the GPS information: 2017-18, 2014, 2011, 2007, and 2004.

To protect participant confidentiality, DHS displaces urban PSUs by up to 2km, rural PSUs by up to 5km, and by up to 10km additionally for 1% of PSUs.^24^ All data from the DHS used in this analysis are publicly available upon request (https://dhsprogram.com/).

Information concerning child immunization is regularly collected in DHS for children under a certain age (three or five years, depending on the survey wave) who are alive at the time of the interview and live with their mothers. For comparability purposes, we focus on children under three years. Usually, a health card is used to determine the immunization status of the child. If a health card is not available or the information on the card is incomplete, the mother’s recall is used instead. We retrieved information about each dose of the four essential childhood vaccines recommended by WHO as part of the basic immunization schedule for young children: three doses of diphtheria–tetanus–pertussis (DTP) vaccine, one dose of bacille Calmette–Guerin (BCG) vaccine, one measles-containing vaccine (MCV), and three doses of oral polio vaccine (OPV)^25^.

The Global Flood Database (GFD) is a database of satellite maps depicting major flooding events that have occurred across the globe since 2000. The database has maps of 913 flooding events available for 169 countries. The images are created by NASA’s MODIS and Terra satellites and are publicly available for download (http://global-flood-database.cloudtostreet.ai/). For each flood event reported, there are five bands of information provided, including the extent of flood water during an event and the duration of the flood in days.^9^ Our analysis includes flood data from seven of the largest floods that occurred in Bangladesh since 2000. These flooding events occurred in 2002, 2003, 2004, 2007, 2010, 2010, and 2017. All data from the GFD used in this analysis are publicly available.

### Exposure

Using a dataset previously compiled by Rerolle et al.^10^, we first constructed a variable indicating whether an area was flood-prone. To determine this, we created a variable representing the percentage of days a given pixel (250m^2^-pixel-level) was classified as flooded out of the total number of days recorded across the seven floods included in the flooding dataset. Then we applied a 10km buffer around the PSUs to account for their displacement and calculated the mean percentage of days flooded in this new buffered area. This information was then used to distinguish between exposed and non-exposed PSUs – any PSU where over five percent of the days were flooded out of the total flood days was considered a flood-prone area and any PSU where less than five percent of the days were flooded was considered not flood-prone. Sensitivity analyses were conducted to explore any differences due to changes in this threshold.^11^

### Outcome

We determine the immunization status of children based on the four vaccines included in the EPI schedule. Usually, BCG is administered shortly after birth, DTP 1–3 and OPV 1–3 are administered at 6, 10, and 14 weeks after birth, and MCV is administered after 9 months of age. We determine whether the child had received every dose of the four recommended vaccines at the time of the interview. To allow for some delay in vaccination, we restrict our sample to children between 12 and 36 months of age. Children who were missing any dose of the four essential vaccines were classified as “non-fully immunized children” (non-FIC). This variable was used in the analysis to examine the relationship between residing in a flood-prone area and not receiving full immunization. More information on the vaccine schedule is summarized in the supplementary materials (Table S1).

### Covariates

We included information on several variables previously shown in the literature to be associated with vaccination status^14,26^. These include age of the mother, household wealth status, education level of the mother, sex of the child, birth order, and residency in a rural or urban area.

### Statistical analysis

As a first step in our analysis, we mapped vaccination coverage across Bangladesh. The geographical distribution of flood-prone areas was also mapped using the Global Flood Database raster data.^11^ To test the associations between living in a flood-prone area and immunization status, we used modified Poisson regressions with robust variance and generalized estimating equations which accounted for correlation at the PSU level. This allowed for the estimation of prevalence ratios (PR).^27^ A univariable analysis was performed as well as an inverse probability of treatment weighting (IPTW) analysis where we checked the covariate balance and accounted for potential confounding^28^. Covariate balance was checked visually with Love plots. To test for effect measure modification on the multiplicative scale, we added an interaction term between the flood-prone variable and urbanity, survey year, birth order, education level, and wealth in our regression models.

### Sensitivity analyses

We first redefined the flooding exposure variable by isolating only the areas that were touched by floods and splitting the PSUs into quartile groups according to the percentage of days that each area was flooded. We then used these quartile groups to construct an ordinal exposure variable, which allowed us to assess a possible dose-response relationship. Additionally, we modified the exposure definition by adjusting the threshold for the percentage of flooded days required for a PSU to be classified as flood-prone. In particular, we applied thresholds of 2% and 10% flooded days to observe how the results vary with a more or less stringent threshold. We also revised the exposure data by removing the 10km buffer area around the PSUs, which was added to account for the PSU displacement in DHS. In this sensitivity analysis, areas were considered flood-prone if the percentage of days flooded was above zero.

We also ran a sensitivity analysis whereby we stratified the sample by age groups of 12-23 months and 24-35 months to examine the possible differences between age groups.^29^ Additionally, we ran a robustness analysis by including only children with vaccine information from vaccination cards and excluding those whose data relied on recall from the mother, which could be subject to recall bias.

All data cleaning, mapping, and analyses were carried out using R software version 4.2.3. All codes needed to reproduce this analysis are available here: https://github.com/EmilieSchwarz/floods_vaccination

## Results

### Descriptive results

Our sample included 19,695 children aged between 0 and 36 months. Of these children, 12,456 had information from their vaccination cards, while 7,239 relied on a mother’s recall. Our sample of children over 12 months old was 13,630 children. Most of the mothers included in our sample had a primary or secondary level of education, and the majority lived in rural areas. When comparing survey waves, we see an increase in the proportion of children who are fully immunized over time (Table 1). A similar trend was observed for each vaccine as well. Overall, 84% of children over 12 months of age in our study sample were classified as fully immunized. The vaccine coverage varies by region, with parts of northeastern and southern Bangladesh displaying lower vaccination rates on average across survey waves compared to other areas (Figure 1). In our sample, we estimate that 17% of children over the age of 12 months live in an area classified as susceptible to flooding. This number varies from 14% to 20% across survey waves. We observed that among children who are not fully immunized, a higher proportion have mothers with no or only primary education, belong to poorer wealth categories, reside in rural areas, and live in flood-prone areas compared to fully immunized children (Table 2).

**Table 1:** Sample descriptive statistics by DHS wave calculated with survey wights. BCG, bacille Calmette–Guerin; DTP, diphtheria–tetanus–pertussis; MCV, measles-containing vaccine; OPV, oral polio vaccine; FIC, fully immunized child; FIC nm, fully immunized child excluding the MCV vaccine.

| Characteristic | Overall N =<br>13,583<sup>1</sup> | 2004 N =<br>2,454<sup>1</sup> | 2007 N =<br>2,166<sup>1</sup> | 2011 N =<br>2,928<sup>1</sup> | 2014 N =<br>2,965<sup>1</sup> | 2017 N =<br>3,070<sup>1</sup> |
| --- | --- | --- | --- | --- | --- | --- |
| **Age of mother** | 25 | 25 | 25 | 25 | 25 | 26 |
| **Education level** |  |  |  |  |  |  |
| no education | 19% | 36% | 25% | 18% | 15% | 6.9% |
| primary | 30% | 32% | 32% | 30% | 28% | 29% |
| secondary | 41% | 26% | 35% | 42% | 47% | 48% |
| higher | 10% | 6.3% | 7.8% | 9.2% | 10% | 17% |
| **Wealth** |  |  |  |  |  |  |
| poorest | 22% | 24% | 20% | 22% | 22% | 22% |
| poorer | 19% | 20% | 19% | 19% | 18% | 20% |
| middle | 18% | 19% | 18% | 18% | 19% | 18% |
| richer | 19% | 16% | 18% | 19% | 21% | 20% |
| richest | 21% | 21% | 24% | 22% | 20% | 21% |
| **Residence** |  |  |  |  |  |  |
| rural | 67% | 69% | 64% | 67% | 68% | 65% |
| urban | 33% | 31% | 36% | 33% | 32% | 35% |
| **Sex of child** |  |  |  |  |  |  |
| female | 49% | 50% | 50% | 50% | 49% | 48% |
| male | 51% | 50% | 50% | 50% | 51% | 52% |
| **Birth order** | 2 (1, 3) | 2 (1, 4) | 2 (1, 3) | 2 (1, 3) | 2 (1, 3) | 2 (1, 3) |
| **Flood-prone area** | 16% | 16% | 13% | 15% | 16% | 18% |
| **BCG** | 97% | 94% | 96% | 97% | 97% | 98% |
| **DTP** | 91% | 83% | 90% | 93% | 92% | 96% |
| **OPV** | 91% | 84% | 91% | 93% | 92% | 95% |
| **MCV** | 86% | 79% | 85% | 88% | 87% | 91% |
| **FIC** | 84% | 76% | 84% | 87% | 85% | 89% |
| **FICnm** | 90% | 82% | 90% | 92% | 91% | 95% |

<sup>1</sup>Sample size; %; Median (Q1, Q3)

**Table 2:** Sample descriptive statistics by immunization status. FIC, fully-immunized child.

| **Characteristic** | **Immunization Status** |  |
| --- | --- | --- |
|  | 0-Non fully immunized,<br>N = 2,111<sup>1</sup> | 1-Fully immunized,<br>N = 11,397<sup>1</sup> |
| **Age of mother** | 25 | 24 |
| **Education level** |  |  |
| no education | 36% | 16% |
| primary | 35% | 29% |
| secondary | 26% | 43% |
| higher | 3.0% | 12% |
| **Wealth** |  |  |
| poorest | 34% | 20% |
| poorer | 21% | 19% |
| middle | 18% | 19% |
| richer | 17% | 19% |
| richest | 10% | 23% |
| **Residence** |  |  |
| rural | 72% | 66% |
| urban | 28% | 34% |
| **Sex of child** |  |  |
| female | 52% | 49% |
| male | 48% | 51% |
| **Birth order number** | 2 | 2 |
| **Flood-prone area** | 21% | 15% |

**Figure 1:**
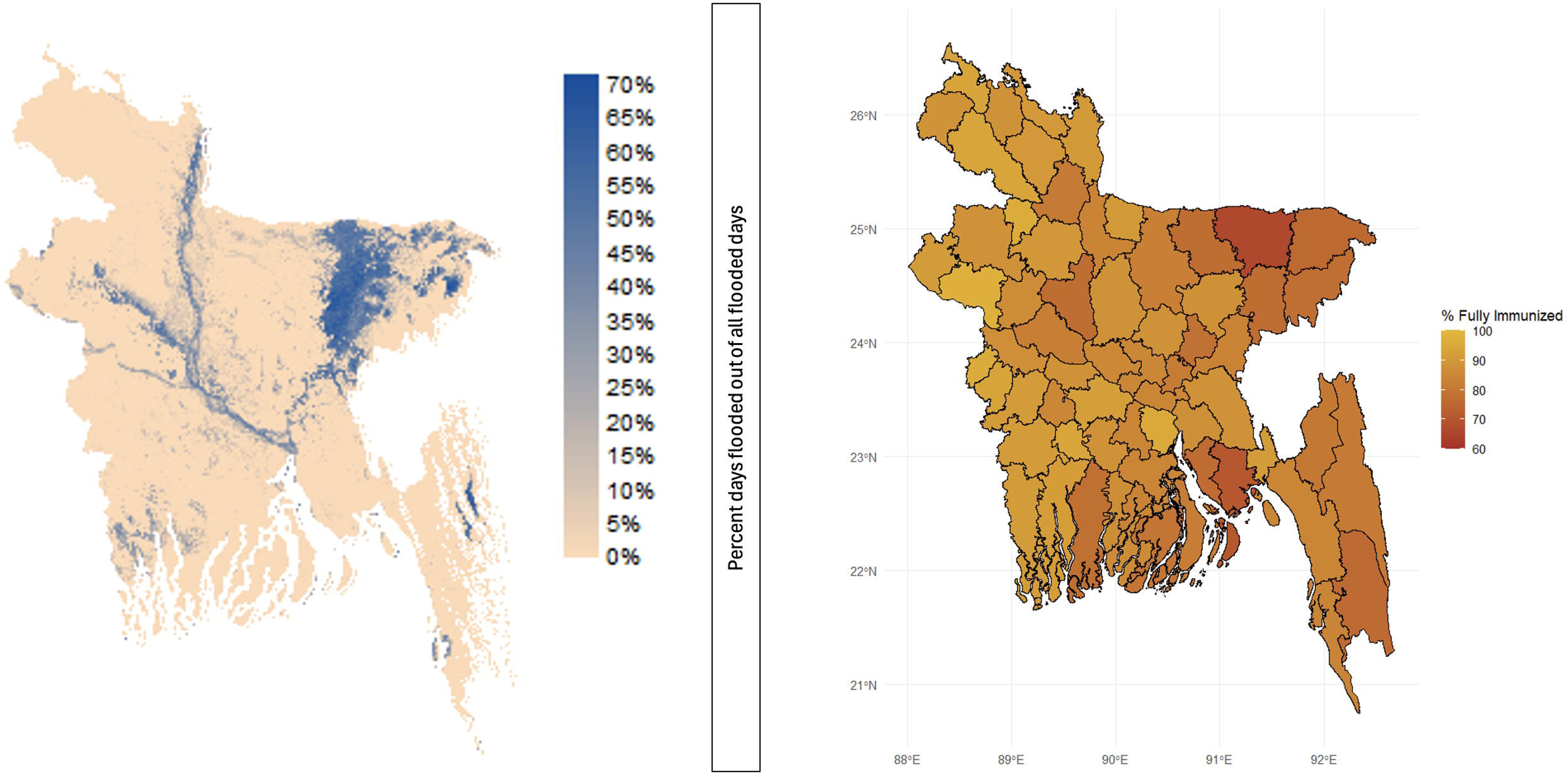
Severity of flooding across Bangladesh (left) and proportion fully vaccinated children by subnational region (right). Flooded days are expressed as the percentage of days flooded out of the total number of flooded days. On the right panel, subnational data are presented at administrative level 2.

### Flood-prone regions and vaccination coverage

The results of the univariable modified Poisson GEE regression suggest that there is a positive relationship between living in a flood-prone area and having an incomplete vaccination status (PR=1.45, CI:1.21-1.74). The direction of the relationship remained but the magnitude of the effect became insignificant after including IPTW considering the confounding role of the age of the mother, education level of the mother, the wealth category of the household, urban and rural residence, sex, and birth order of the child (PR=1.05, CI:0.90-1.22). The covariate balance and weight distribution resulting from the IPTW showed a minimal difference between exposed and unexposed across all considered covariates (see Figure S3).

When exploring the relationship with individual vaccines, the results suggest that there is a positive relationship between living in a flood-prone area and incomplete vaccination for DTP (PR = 1.24, CI: 1.05-1.46), BCG (PR = 1.27, 1.08-1.50), and OPV (PR = 1.24, CI: 1.06-1.46). The direction of the relationship remained but the effect became non-significant when looking at the MCV vaccine alone (PR = 1.06, CI: 0.90-1.25). When looking at full vaccination while excluding Measles, the relationship remained positive and statistically significant (PR = 1.23, CI: 1.05-1.44). (Figure 2).

The positive relationship observed for the DTP, BCG, and OPV vaccines persisted across various sensitivity analyses, although the effect size varied in magnitude (Figures S1 and S2). Specifically, when excluding children with missing vaccination records, the positive association remained, and the effect size increased for DTP, BCG, and OPV (Figure S1). The direction of the association remained consistent across different age groups, though the magnitude varied (Figure S1). Additionally, the association remained significant for BCG, DTP, and OPV when different exposure thresholds were applied, with significant findings for 10%, 2%, and >0% flooding exposure. However, the analysis based on flooding quartiles resulted in smaller sample sizes and lacked sufficient power to detect any meaningful relationship (Figure S2).

We further investigated effect measure modification by urban-rural residence. This analysis revealed that the relationship was observed in rural areas but not in urban areas, across most vaccines and doses (Figure S1 and S2). The only exception was the second dose of the DTP vaccine, for which the association was statistically significant in both rural and urban populations. Additionally, the relationship for the measles vaccine, as well as for all vaccines combined, was not statistically significant in either rural or urban populations.

**Figure 2.**
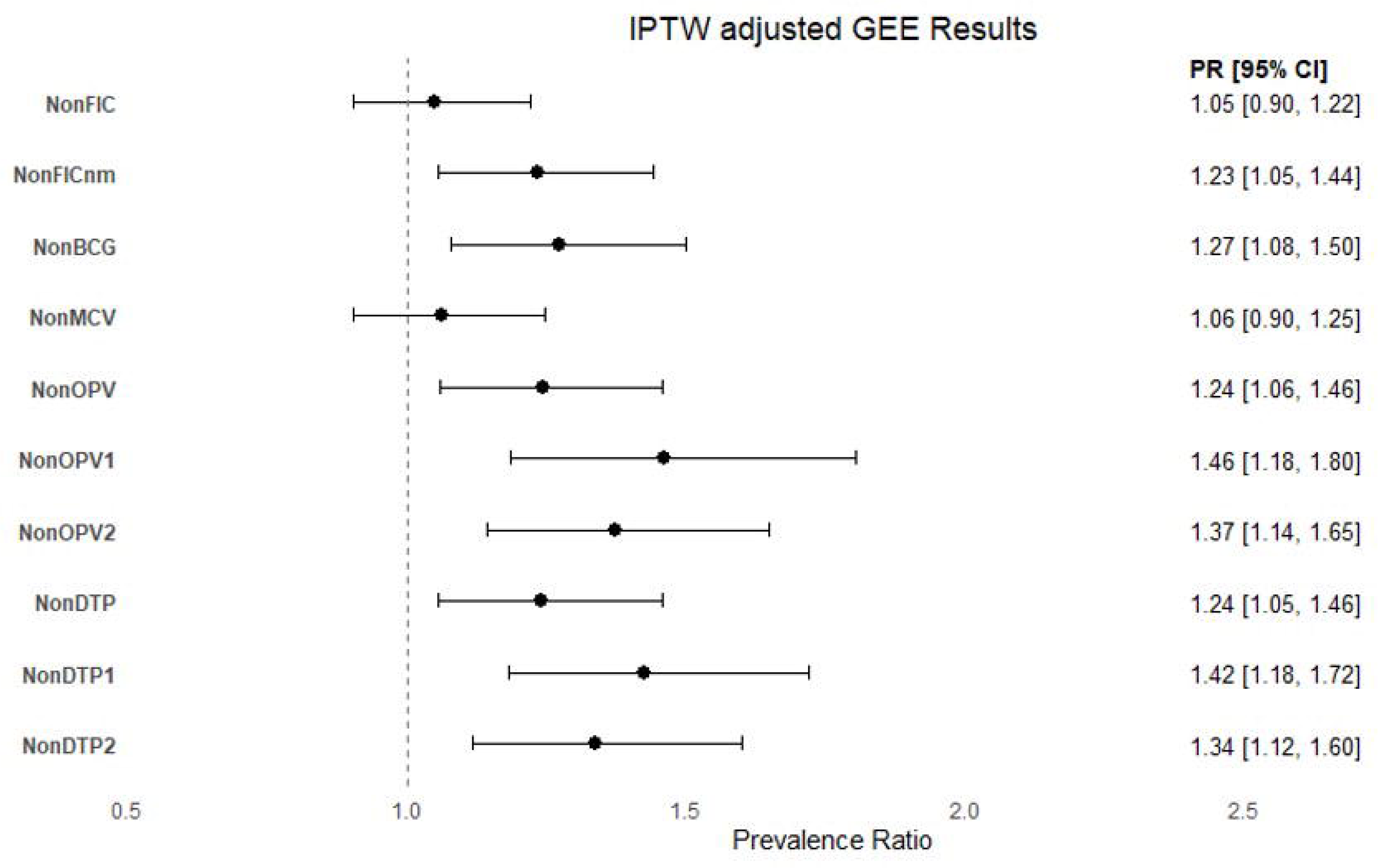
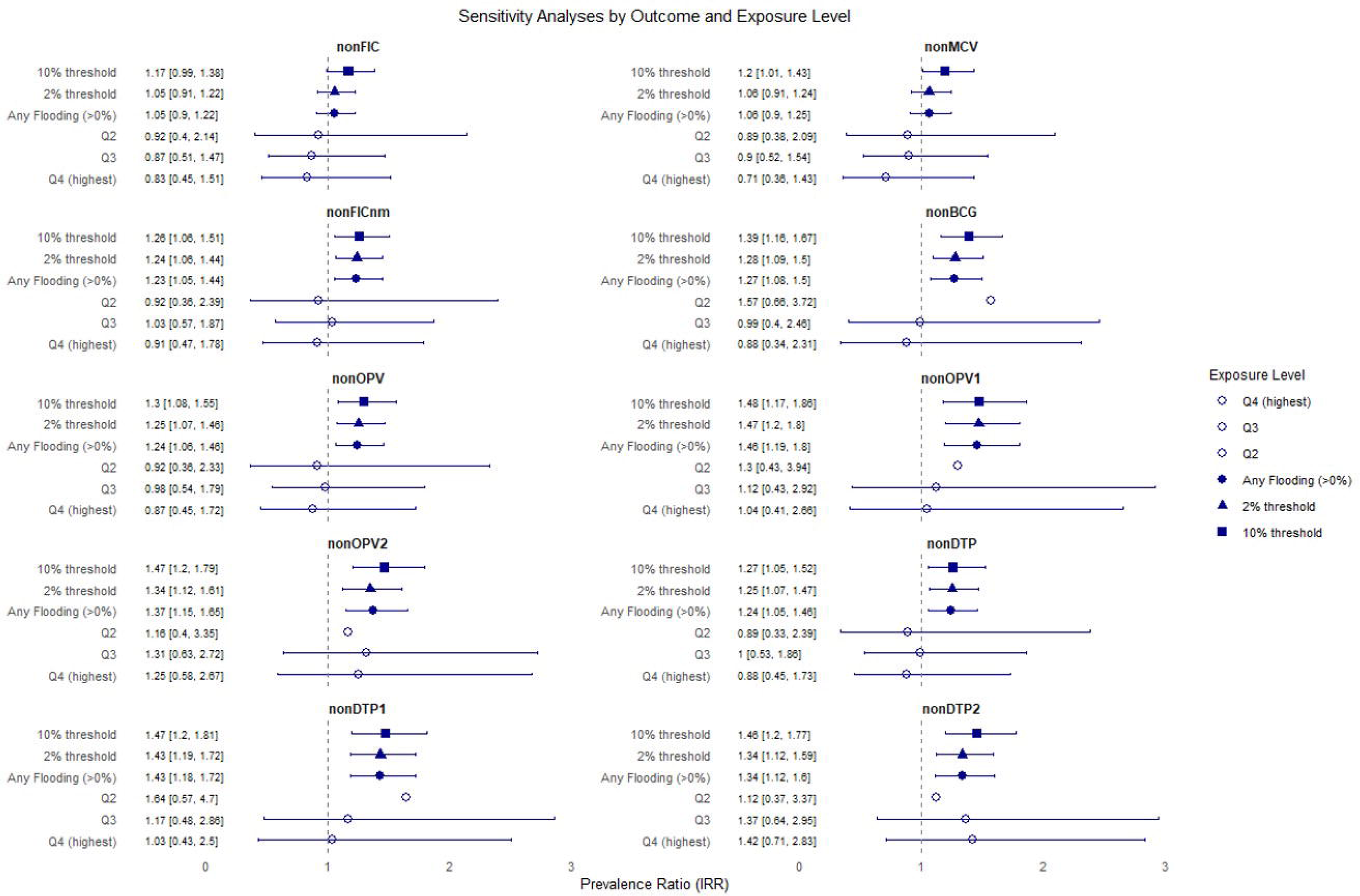
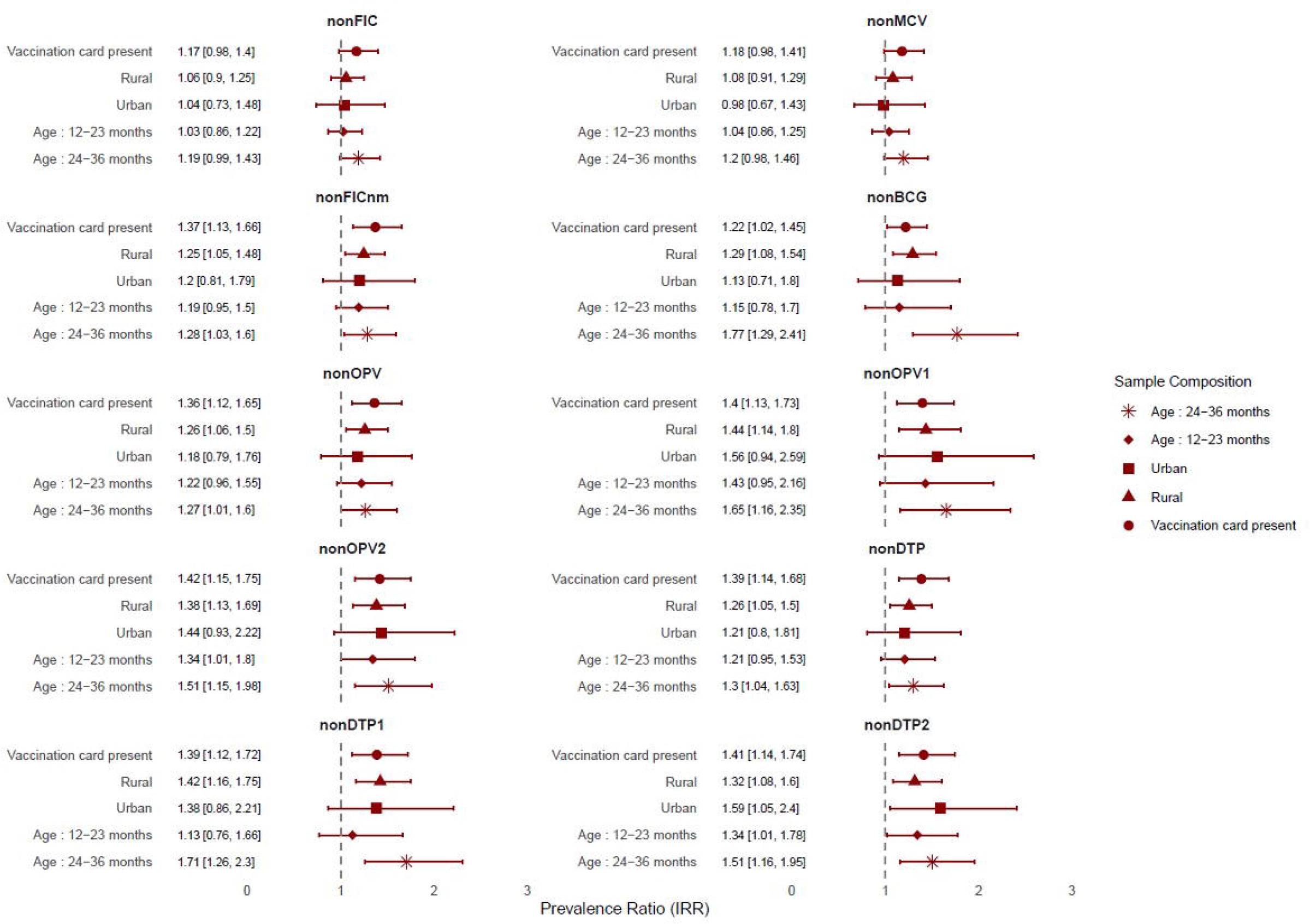
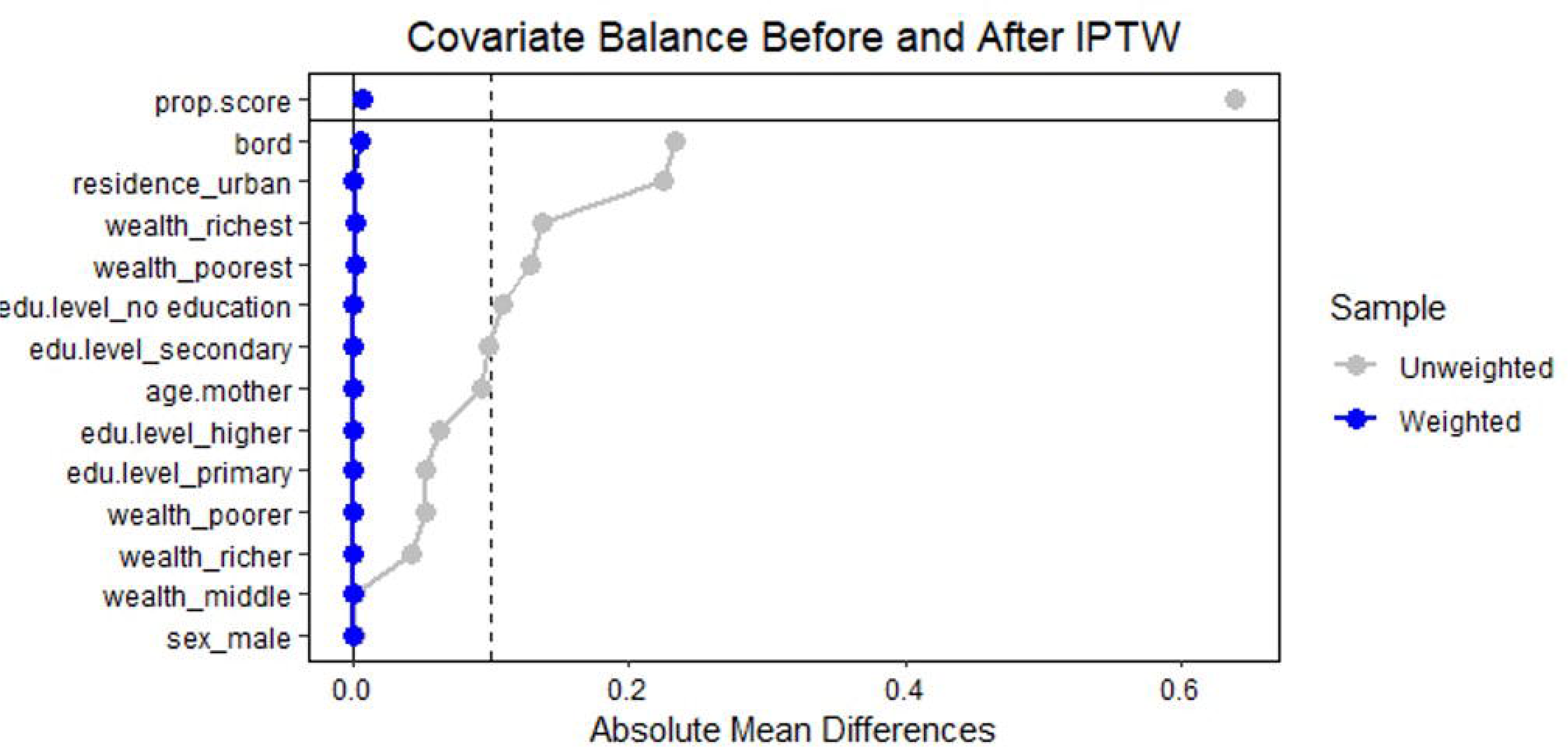
Association between living in a flood-prone area and incomplete vaccination status-IPTW results across vaccine types and doses.

## Discussion

The findings of this study suggest that residing in flood-prone areas is associated with an increased likelihood of incomplete childhood vaccination for DTP, BCG, and OPV in Bangladesh. These results remain robust even after accounting for geographical factors, controlling for potential confounders, and conducting several sensitivity analyses. However, no significant association was observed for the measles vaccine or for the full vaccination coverage. This may indicate that measles-specific vaccination campaigns in Bangladesh have been particularly effective. Additionally, since the measles vaccine is administered at a later age, its inclusion in the overall vaccination analysis changes the population, resulting in a smaller sample size which may have limited statistical power.

To our knowledge, no previous research has explored quantitatively the impact of floods on vaccination. Our findings are consistent with the broader literature, which suggests that populations exposed to floods may experience interruptions in healthcare services due to infrastructure and road damage, supply shortages, and forced relocation.^4,31^ For instance, a 2008 study documented how floods damaged the infrastructure of schools, homes, businesses, and health facilities across Mozambique, Zambia, Zimbabwe, and Malawi^4^. These floods had notable impacts on public health, leading to thousands of reported cholera cases in the subsequent months^4^. Earlier research has also described how flooding may specifically disrupt vaccine supply chains due to transportation capacity issues and potential cold-chain breakdowns.^31^

Vaccination is widely recognized as a critically important global public health intervention. Disruptions in routine childhood vaccination systems can result in numerous excess deaths globally. Identifying and understanding these risks is crucial for safeguarding population health. The findings of our study indicate that a significant number of children lack immunizations due to residing in flood-prone areas. Our results attest to the magnitude of this issue and its potential ramifications for public health. The incomplete vaccination status of children increases their susceptibility to contracting preventable infectious diseases and this risk may be amplified during extreme climate conditions. For example, a recent study showed that precipitation anomalies are associated with an increased risk of diarrheal diseases in young children.^32^ As climate change impacts become more pronounced, it is increasingly important that the disaster management systems set in place integrate public health needs. Understanding the vulnerabilities that are exacerbated during flooding events could inform national response priorities and facilitate the prevention of health emergencies during extreme climate events.

Populations in flood-affected areas are likely to experience disruptions in other routine healthcare services, such as health checks, screenings, or counseling. To safeguard population health, policymakers could implement several targeted measures. Firstly, healthcare access tends to be disrupted during floods due to infrastructure damage, such as collapsed roads and buildings. To address this issue, the infrastructure in such vulnerable areas should be reinforced to ensure uninterrupted access to care during floods and other climate emergencies. Secondly, catch-up vaccination campaigns could be launched once access to the flood-affected areas has been improved. During the COVID-19 pandemic, such disruptions in routine vaccination were experienced across the world. To prevent the long-term impacts of these disruptions, catch-up vaccination efforts were put into place^33^. Similarly, catch-up vaccination campaigns could be launched in flood-affected areas to prevent immunization gaps.

Our study has a few limitations that need to be considered. Firstly, the spatial displacement of PSU locations in DHS means that the geographical estimates are not completely accurate. We accounted for this by adding a buffer around the PSUs and running sensitivity analyses on the exposure definition but we cannot rule out potential exposure misclassification. Nevertheless, such misclassification would be non-differential and would likely lead to an underestimation of the true effect size. Another limitation concerns the exposure variable, which relies on geographical data from seven floods in Bangladesh, assuming these floods are representative of the country’s flood-prone areas. This assumption is considered reasonable due to the consistent flooding patterns observed across the selected events, which was demonstrated in an earlier analysis in which researchers studied the association between residing in flood-prone areas and infant mortality.^11^ It is also important to acknowledge the potential presence of bias in the vaccination data. Recall bias may arise when vaccination records are unavailable, requiring reliance on maternal recall to determine if a child received all vaccine doses. Additionally, social desirability bias could affect reporting accuracy, as mothers might alter their responses if they believe the interviewer disapproves of their child’s vaccination status. To account for such bias, we conducted robustness analyses by excluding cases prone to it, and the association remained unchanged. Finally, our results may be subject to survivor bias, given that some of the children who lacked vaccination may have died before the time of the interview. If this is the case, our sample would include fewer children who were exposed and unvaccinated, suggesting that our estimates are biased downwards. Despite these limitations, our results remain consistent across various robustness analyses, and there is a possibility that we are underestimating the true association.

This work suggests that an association exists between residing in flood-prone areas and missing certain vaccines. Future work can explore specific factors that exacerbate or mitigate the impact of floods on essential childhood immunization. For example, the distance to a health facility may be an important determinant of general access to immunization services. Addressing inequalities in access to vaccination will be critical for closing vaccine gaps across and within countries.^15^ Parental opinions on vaccination can also influence child immunization rates. This information, gathered in recent DHS rounds since the onset of the COVID-19 pandemic, could be valuable for future research. Understanding the effectiveness of preventive education and structural reinforcement programs in shaping parental attitudes will be crucial for optimizing public health resources. Religious beliefs also have been shown to play a role in vaccination status and could be investigated in this context, given that Bangladesh has four main religious groups^34^. In the case of a large discrepancy in vaccination rates across population subgroups, community engagement efforts should be reinforced to encourage vaccination among under-vaccinated communities.

This research utilized vaccination data collected between 2004 and 2019, thus focusing on the pre-pandemic period. A growing body of literature shows that the pandemic had important implications for vaccination worldwide. In particular, research has shown that routine immunization during the COVID-19 pandemic was disrupted in high-income countries and LMICs alike, but more severe disruptions were observed in LMICs.^35^ Considering that Bangladesh suffered severe immunization disruptions during the COVID-19 pandemic, it would be important to understand how flooding may have posed additional challenges in terms of access and utilization of routine immunization services during the pandemic period.

Bangladesh is facing disproportionate impacts from the effects of climate change in comparison to its minimal contributions to global greenhouse gas emissions. While some of the direct impacts of climate change on human health have been well documented, the indirect impacts on health are not yet well understood. This work is a step in this direction. Understanding the myriad ways in which climate change impacts human health will be critical for developing appropriate prevention and intervention strategies that safeguard population health in the presence of more frequent and extreme weather events.

## Data Availability

All codes to reproduce the analysis are available in the manuscript and all data is publically available upon request.

https://global-flood-database.cloudtostreet.ai/

https://dhsprogram.com/countries/Country-Main.cfm?ctry_id=1

