## Supplementary Files for "Association Between Residence in Flood-Prone Areas and Incomplete Childhood Vaccination Coverage in Bangladesh"

**Table S1. Vaccine Administration Timing and Ages Included in Analysis**

| **Vaccine** | **Recommended timing** | **Ages included in the analysis** |
| --- | --- | --- |
| Bacillus Calmette-Guérin | At birth | All ages |
| Diptheria- tetanus-pertussis |  |  |
| *Dose 1* | 6 weeks | 2 months and older |
| *Dose 2* | 10 weeks | 4 months and older |
| *Dose 3 (all)* | 14 weeks | 6 months and older |
| Polio |  |  |
| *Dose 1* | 6 weeks | 2 months and older |
| *Dose 2* | 10 weeks | 4 months and older |
| *Dose 3 (all)* | 14 weeks | 6 months and older |
| Measles | 9 months | 12 months and older |

**Figure S1. Association between living in a flood-prone area and incomplete vaccination status across various exposure definitions**

[Fig-S1.jpeg]

**Figure S2. Association between living in a flood-prone area and incomplete vaccination status across various population definitions**

[Fig-S2.jpeg]

**Figure S3. Covariate balance love plot**

[Fig-S3.jpeg]
